# The Burden of Generational Harm due to Alcohol use in Tanzania: a mixed method study of pregnant women

**DOI:** 10.1101/2024.08.22.24312125

**Authors:** Kirstin West, Alena Pauley, Mia Buono, Miriana Mikindo, Yvonne Sawe, Joseph Kilasara, Francis Sakita, Sharla Rent, Bariki Mchome, Blandina T. Mmbaga, Catherine A. Staton

**Affiliations:** Duke Global Health Institute, Duke University, Durham, NC USA; Kilimanjaro Clinical Research Institute, Moshi, Tanzania; Department of Clinical Nursing, Kilimanjaro Christian Medical University College, Moshi, Tanzania; Kilimanjaro Christian Medical Center, Moshi, Tanzania; Kilimanjaro Christian Medical University College, Moshi, Tanzania; Duke Department of Pediatrics, Duke University School of Medicine, Durham, NC USA; Department of Obstetrics and Gynecology, Kilimanjaro Christian Medical Center, Moshi, Tanzania; Department of Emergency Medicine, Duke University School of Medicine, Durham, NC USA

**Keywords:** pregnancy, alcohol use, Tanzania, mixed-methods

## Abstract

**Background:** Rates of prenatal alcohol use in Sub-Saharan Africa (SSA) are increasing, despite regulatory bodies urging pregnant women to abstain from alcohol. Tanzania has minimal policies, interventions, and young female education addressing alcohol consumption during pregnancy (ACDP), leading to a considerable number of pregnancies being exposed to alcohol and consequent health consequences like fetal alcohol spectrum disorder (FASD). Research investigating the prevalence of ACDP in SSA —specifically in Tanzania— is abundant. In Tanzania, there is a limited understanding of alcohol use practices among pregnant women who consume alcohol (PWCA), as well as community knowledge, attitudes, and cultural beliefs related to ACDP.

**Methods:** A total of 655 individuals were enrolled in this sequential, explanatory mixed-methods study using systematic random sampling between October 2021 and May 2022. Quantitative survey data from 533 female patients presenting for care at KCMC ED or RHC were analyzed to compare sociodemographic factors and alcohol use practices among pregnant, younger non-pregnant, and older non-pregnant women using descriptive frequencies in RStudio. Nineteen participants were purposively selected based on quantitative survey data for qualitative semi-structured IDIs exploring knowledge, attitudes, and cultural beliefs surrounding ACDP. A grounded theory approach was used to analyze in-depth interviews (IDIs) in NVivo.

**Results:** A large percent of pregnant women reported alcohol consumption of at least once per week (42.2%). Older non-pregnant women had the highest rates of alcohol use per week (66.0%) and were more likely to believe alcohol use was acceptable during pregnancy. Younger non-pregnant women reported the highest weekly alcohol expenses and held the highest prevalence of harmful or hazardous drinking (HHD) (16.4%). Average [SD] AUDIT scores were 1.70 [3.28] for pregnant women, 2.94 [4.79] for younger non-pregnant women, and 2.51 [4.36] for older non-pregnant women. Older non-pregnant women exhibited the highest prevalence of depression (31.4%). Average [SD] PHQ-9 scores were 4.71 [3.12] for pregnant women, 5.85 [4.80] for younger non-pregnant women, and 7.29 [5.55] for older non-pregnant women. Qualitative analyses demonstrated that (1) cultural beliefs are intricately tied to perceived benefits of ACDP, (2) a history of alcohol use preceding pregnancy largely influences ACDP, and (3) community views of PWCA are negative.

**Significance:** Our findings demonstrate a concerning trend of ACDP in Moshi, Tanzania. Cultural and community beliefs, along with limited knowledge of ACDP, among women of all ages appear to influence ACDP and community views of PWCA. Consequently, community-wide education initiatives and pre-pregnancy interventions highlighting the dangers of ACDP are necessary. Holistic support services may help curb alcohol use and improve birth experiences. Further research is needed to explore ACDP as a form of self-medication for depression, fear, anxiety, and pain in pregnant women in SSA.

## 1 INTRODUCTION

As alcohol use increases globally, so do the number of alcohol-related health consequences, including death and disability ^1,2^. In a 2020 global risk assessment, alcohol consumption ranked among the top ten factors impacting overall health, and served as a major risk factor for communicable, maternal, perinatal, and nutritional diseases ^2^.

A notable, well-established alcohol-related harm is the detrimental effect on pregnancy ^2–4^. Alcohol use during this time period can cause an increased risk of birth defects, developmental disabilities, miscarriage, intrauterine growth restriction, spontaneous abortion, preterm labor, and stillbirth ^3,4^. Global estimates from 1984 to 2014 indicated that approximately 9.8% of pregnant women worldwide consumed alcohol^5^. Particularly prevalent in low- and middle-income countries, rates of alcohol consumption during pregnancy (ACDP) can range from 2.5% in Nigeria to as high as 32.5% in the Democratic Republic of the Congo (Popova et al., 2016). Fetal alcohol spectrum disorder (FASD) —a congenital syndrome caused by intrauterine fetal alcohol expose and characterized by growth retardation, facial dysmorphology, cognitive dysfunction, and neurobehavioral disabilities in the fetus— has an estimated prevalence of 14.8% in Africa, further highlighting alarming rates of ACDP in the region ^5,7,8^. The WHO African region was found to have the highest prevalence of binge drinking, defined as having four or more drinks on a single occasion, during pregnancy, estimated at 3.1% ^9^. This is increasingly concerning as populations with high rates of binge drinking have been linked to similarly high rates of FASD ^10–12^.

Existing literature emphasizes the profound health consequences of ACDP in sub-Saharan Africa (SSA), necessitating increased awareness and robust intervention strategies. The World Health Organization (WHO) urges pregnant women to abstain from alcohol use due to the health risks for the mother and fetus ^13^. However, countries such as Tanzania have minimal policy and health system work addressing ACDP. Further, the findings regarding the association between alcohol use and perinatal harm are not always communicated to pregnant women. The lack of intervention and young female education contributes to a considerable number of pregnancies in Tanzania being subject to alcohol exposure, with nearly 20% in Northern Tanzania and 15% in Dodoma, the capital of Tanzania ^14^. In Moshi, Tanzania, rates of alcohol use are 2.5 times higher than the national average, which raises concerns that similar patterns of alcohol consumption likely extend to pregnant women, warranting further investigation^15^.

Cultural practices and beliefs are intricately linked to ACDP in SSA, impacting pregnant women’s decision to alcohol consumption. Cultural traditions and values are passed down through generations and constitute knowledge that effectively shapes pregnant individuals’ alcohol use practices, complicating some education efforts. Comprehending the extent and depth of cultural influences on ACDP is critical to developing culturally appropriate interventions aimed at preventing alcohol use among pregnant women.

Research investigating the prevalence of ACDP in SSA is abundant ^6,9,14,16^. However, there is a limited understanding of alcohol use practices, the knowledge and attitudes related to ACDP, and the ways these impact cultural beliefs among both pregnant women and women of child-bearing age. This information is needed to inform future intervention design and policy efforts, as well as bolstering education efforts for women of childbearing age, with the ultimate goal of better supporting pregnant women and reducing alcohol-related pregnancy complications. This study aims to address this gap by providing a more comprehensive understanding of the sociocultural factors, such as knowledge and attitudes, influencing alcohol use practices among pregnant women presenting to either the Kilimanjaro Christian Medical Centre (KCMC) Emergency Department (ED) or Reproductive Health Center (RHC). The objectives of this study are to (1) evaluate the proportion of pregnant women who consume alcohol (PWCA), (2) investigate consumption patterns and alcohol use practices of pregnant women and non-pregnant women, accounting for child-bearing age (3) assess community knowledge, attitudes, and cultural beliefs on ACDP.

## 2 METHODS

### Study Design Overview

This analysis originates from a sequential, explanatory mixed-methods study conducted in Moshi, Tanzania —an urban town with over 200,000 residents— focused on exploring unhealthy alcohol consumption at the KCMC ED and RHC. The high concentration of injury patients at KCMC’s ED and the large female patient population at the RHC provide ideal settings to study risky alcohol use behaviors, particularly among pregnant women. Participants were prospectively recruited for this study starting in October 2021 and ending in May 2022. Quantitative surveys and qualitative semi-structured in-depth interviews (IDIs) were conducted, with surveys used to guide purposeful qualitative data sampling. A total of 541 women were enrolled in this study, with 533 providing complete survey data. In addition to women, men were included in in-depth interviews (IDIs) to provide a broader community perspective on ACDP, totaling 10 female and 9 male IDI participants. A thorough overview of the methods has been previously published ^17^.

### Exclusion Criteria

Those eligible for study participation had sought medical care at the KCMC ED or the KCMC RHC^18^, were over the age of 18, not prisoners, fluent in Kiswahili, and able to provide informed consent. Ability to provide consent was defined as being clinically sober, medically stabilized, and physically able to complete the survey verbally. Pregnant females were defined as women who had a confirmed in-utero pregnancy, based on self-report. Literature defines the reproductive lifespan of women as 45 years and below^45^. This criterion was used as the cut-off value to distinguish between childbearing age (≤45 years; e.g., younger) and non-childbearing age (46+ years; e.g., older) among non-pregnant women.

### Quantitative Data

#### Procedures

Using intake triage registries from the ED and the RHC, a systematic random sampling approach was used to approach every third patient seeking care. Gender-matched Tanzanian research assistants proficient in Kiswahili provided a comprehensive overview of the study to participants in a quiet and private clinic space, only after the patient had achieved medically stability and clinical sobriety. Patients were given the opportunity to inquire about the study, decline participation, or express interest. For those willing to participate, a detailed discussion regarding the study, including potential risks and benefits, took place. Written informed consent was then obtained. Patients who were illiterate marked their initials or a cross mark. Survey questions were read aloud to encourage participation across different literacy levels.

#### Instruments

Quantitative surveys were the primary instruments, assessing baseline sociodemographic factors, self-reported alcohol use, Alcohol Use Disorder Identification Test (AUDIT) scores, Drinker’s Inventory of Consequences (DrInC) scores, and Patient Health Questionnaire (PHQ-9) depression scores. The Tanzanian research team translated, reviewed, revised, and pilot-tested all survey questions on sociodemographic data and self-reported alcohol use prior to data collection. Participants were asked to report their alcohol consumption quantity in terms of standard drinks, per WHO guidelines where one standard drink contains 10 grams of pure ethanol ^19^.

To comprehensively assess alcohol consumption and unhealthy alcohol-related behaviors among pregnant women, the AUDIT survey tool —with scores ranging from 0 to 40— was employed ^20,21^. An AUDIT score of 8 or above indicates clinically significant harmful or hazardous drinking (HHD), in both global and study settings ^20,22–25^. Patients with HHD (AUDIT ≥ 8) were identified, as their alcohol consumption places them at risk for serious health consequences, necessitating additional alcohol-related treatment and social support ^26^.

The DrInC survey quantitatively evaluates alcohol-related consequences, with higher scores indicating greater consequences. It includes 50 yes-or-no questions across five domains: physical, intrapersonal, social responsibility, interpersonal, and impulse control ^27–29^. The DrInC tool does not measure severity or frequency of alcohol-related consequences, and no clinically relevant cut-off score exists. This survey was cross-culturally adapted and clinically tested at KCMC before use in this study ^29^.

The PHQ-9 tool is globally recognized for diagnosing depression, scaled from 0 to 27. It includes 9 items assessing depressive symptoms such as appetite changes, sleep patterns, and thoughts of self-harm, with responses to each item scored from 0 to 3 ^30–32^. Higher scores indicate more severe depression. Translated into Kiswahili and validated in Tanzania, a score of 9 is the optimal cut-off for identifying major depressive episodes (MDD), classifying patients into depression (PHQ-9 ≥ 9) and non-depression (PHQ-9 < 9) groups ^30,31,33–36^.

#### Analysis

Descriptive frequencies and statistics were used to compare sociodemographic factors and alcohol use between pregnant women, non-pregnant women of childbearing age (≤45 years), and non-pregnant women not of childbearing age (>45 years). Quantitative measures, including AUDIT, DrInC, and PHQ-9 scores, were also compared between pregnant and non-pregnant women. Continuous and categorical data were compared using Analysis of Variance (ANOVA) type 3 sum of squares, and Pearson’s chi-square test, respectively. Data analyses were performed using user-tested packages in RStudio (version 1.4).

### Qualitative Data

#### Sampling

Individuals from the quantitative survey were purposefully sampled to participate in semi-structured IDIs, ensuring a diverse range of demographic backgrounds like age, marital status, education level, occupation, tribe, and religion. Those considered for IDIs were individuals reporting neutral or positive past experiences with alcohol, former alcohol users who are currently abstinent, patients with close friends or family members who consume alcohol heavily, and those who suffered adverse consequences from their own drinking. This methodological approach aimed to capture a wide range of perspectives on ACDP. To further ensure a holistic view of community knowledge and personal perspectives on ACDP, a gender-balanced approach was used. That is, we aimed to interview ten males and ten females since data saturation is typically reached around 20 interviews ^37–39^. However, saturation was reached with 19 IDIs, comprising nine males and ten females

#### Procedures

Upon completion of the quantitative survey, research assistants identified suitable candidates for IDIs using their professional judgment, with monthly reviews of sociodemographic and alcohol-use-related data conducted to ensure diverse representation of participants. Consenting participants provided their phone numbers to schedule interviews.

Interviewers were fluent in Kiswahili, gender-matched, and had previously established rapport with the patients during the survey phase to foster open and candid discussions, especially on sensitive and stigmatized topics. Prior to starting the one-on-one interviews, the study goals were communicated, and participants received 5,000 TSH (∼2 USD) to cover transportation costs. All interviews were audio-recorded, typically lasting 60 to 100 minutes, and included a break with snacks. These semi-structured IDIs were assigned unique identification numbers, ranging from IDI #1 to IDI #19, of which were known only to the research team.

#### Instrument

The interview guide utilized for IDIs included open-ended questions —originally created in English, and then translated into Kiswahili— with probes built in for additional questioning when the subject matter was ambiguous, vague, interesting, or inconsistent. Cultural appropriateness and relevancy were ensured during pilot testing. The standardized interview guide and related probes (Supporting File 1/Appendix) were used for all participants to assess community knowledge, views, and perspectives on ACDP.

#### Analysis

Qualitative data was analyzed using an applied, thematic, grounded theory approach, where IDI responses were collected and analyzed without pre-existing theoretical frameworks ^40,41^. Considering research on pregnant female drinking behavior is limited in Moshi, the grounded theory approach was well-suited to capture emerging themes. The Tanzanian research team was trained on qualitative analysis and interview coding using NVivo 12. An iterative development process using inductive and deductive coding schemes resulted in an initial codebook. To facilitate an ongoing synthesis of data and maintain a dynamic codebook, content memos were created with each emerging theme, and subsequent code. To enhance content validity and cultural accuracy, the final analysis was discussed between the entire research team.

### Ethics Statement

Prior to data collection, ethical approval for this study was obtained from the Duke University Institutional Review Board, the Kilimanjaro Christian Medical University College Ethical Review Board, and the Tanzanian National Institute of Medical Research. Written consent was formally obtained from all participants. Personal health information used during screening and enrollment was de-identified for confidentiality throughout data collection, storage, analysis, and sharing. Data was only shared in accordance with a data sharing agreement.

## 3 RESULTS

### Quantitative

The study sample consisted predominantly of women who self-identified as Christian (81.2%), and were affiliated with the Chagga tribe (50.0%), with ages spanning 17 to 95 years. On average, pregnant women were 29.1 years younger than non-pregnant women, with over half of pregnant women between the ages of 25 to 34 years (Table 1). Notable differences in marital status were also observed between pregnant women and non-pregnant women. For women of childbearing age, those who were more pregnant were more likely to be living with a partner in either a registered (61.2% vs 42.2% in non-pregnant women) or unregistered marriage (29.3% vs 12.9% in non-pregnant women). In comparison, those not pregnant were more likely to be single (39.7% vs 8.6% among pregnant women) or divorced (3.4% vs 0.9% among pregnant women). Younger women (pregnant or non-pregnant) were also more likely to have attained a college-level education (51.7% and 50.8% vs 9.2%), be employed (72.4% and 62.1% vs 47.6%), and have higher personal and household incomes than older, non-childbearing women. These values and others can be seen in Table 1.

**Table 1:** Study Population Demographics.

| **Demographics Stratified by Pregnancy Status** | **Overall**<br>**(n = 533)** | **Pregnant**<br>**(n = 116)** | **Not Pregnant, Child-**<br>**bearing**<br>**(n = 232)** | **Not Pregnant, Not**<br>**Child-bearing**<br>**(n = 185)** | **P Value** |
| --- | --- | --- | --- | --- | --- |
| **Age (in years), missing: 6 pregnant** | 40.9 (16.0) | 30.1 (5.82) | 31.1 (7.78) | 59.4 (9.98) | **<0.001 \*\*\*** |
| **Age Category (in years), missing: 6 pregnant** |  |  |  |  | **<0.001 \*\*\*** |
| 18 - 24 | 78 (14.6%) | 19 (16.4%) | 59 (25.4%) | 0 (0%) |  |
| 25 - 34 | 155 (29.1%) | 67 (57.8%) | 88 (37.9%) | 0 (0%) |  |
| 35 - 44 | 101 (18.9%) | 23 (19.8%) | 78 (33.6%) | 0 (0%) |  |
| 45 - 54 | 79 (14.8%) | 1 (0.9%) | 7 (3.0%) | 71 (38.4%) |  |
| Over 55 | 114 (21.4%) | 0 (0%) | 0 (0%) | 114 (61.6%) |  |
| **Religion, missing: 0** |  |  |  |  | 0.6886 |
| Christian | 433 (81.2%) | 94 (81.0%) | 186 (80.2%) | 153 (82.7%) |  |
| Muslim | 91 (17.1%) | 21 (18.1%) | 43 (18.5%) | 27 (14.6%) |  |
| None | 8 (1.5%) | 1 (0.9%) | 3 (1.3%) | 4 (2.2%) |  |
| Other | 1 (0.2%) | 0 (0%) | 0 (0%) | 1 (0.5%) |  |
| **Marital Status, missing: 0** |  |  |  |  | **<0.001 \*\*\*** |
| Living with a partner in a registered marriage | 263 (49.3%) | 71 (61.2%) | 98 (42.2%) | 94 (50.8%) |  |
| Living with a partner, not in a registered marriage | 66 (12.4%) | 34 (29.3%) | 30 (12.9%) | 2 (1.1%) |  |
| Never Married or Single | 169 (31.7%) | 10 (8.6%) | 92 (39.7%) | 67 (36.2%) |  |
| Divorced or Separate | 23 (4.3%) | 1 (0.9%) | 8 (3.4%) | 14 (7.6%) |  |
| Widowed | 12 (2.3%) | 0 (0%) | 4 (1.7%) | 8 (4.3%) |  |
| **Highest Educational Attainment, missing: 1** |  |  |  |  | **<0.001 \*\*\*** |
| None | 8 (1.5%) | 0 (0%) | 1 (0.4%) | 7 (3.8%) |  |
| Primary | 148 (27.8%) | 7 (6.1%) | 33 (14.3%) | 108 (58.4%) |  |
| Secondary | 131 (24.6%) | 40 (34.5%) | 58 (25.1%) | 33 (17.9%) |  |
| Vocational | 33 (6.2%) | 4 (3.4%) | 14 (6.0%) | 15 (8.1%) |  |
| College | 195 (36.6%) | 60 (51.7%) | 118 (50.8%) | 17 (9.2%) |  |
| Graduate | 17 (3.2%) | 5 (4.3%) | 8 (3.5%) | 4 (2.2%) |  |
| **Employment Status, missing: 0** |  |  |  |  | **<0.001 \*\*\*** |
| Employed | 316 (59.3%) | 84 (72.4%) | 144 (62.1%) | 88 (47.6%) |  |
| Unemployed | 156 (29.3%) | 20 (17.2%) | 41 (17.7%) | 95 (51.4%) |  |
| Student | 61 (11.4%) | 12 (10.3%) | 47 (20.3%) | 2 (1.1%) |  |
| **Tribe, missing: 0** |  |  |  |  | **<0.01 \*\*** |
| Chagga | 267 (50.0%) | 64 (55.2%) | 97 (41.8%) | 106 (57.3%) |  |
| Pare | 50 (9.4%) | 9 (7.8%) | 16 (6.9%) | 25 (13.5%) |  |
| Sukuma | 26 (4.9%) | 8 (6.9%) | 13 (5.6%) | 5 (2.7%) |  |
| Massai | 21 (3.9%) | 5 (4.3%) | 8 (3.4%) | 8 (4.3%) |  |
| Sambaa | 16 (3.0%) | 3 (2.6%) | 9 (3.9%) | 4 (2.2%) |  |
| Nyaturu | 17 (3.2%) | 3 (2.6%) | 13 (5.6%) | 1 (0.5%) |  |
| Other African | 135 (25.3%) | 24 (20.7%) | 76 (32.8%) | 35 (18.9%) |  |
| Non-African | 1 (0.2%) | 0 (0%) | 0 (0%) | 1 (0.5%) |  |
| **Personal Income (in TZS), missing: 3** |  |  |  |  | **<0.001 \*\*\*** |
| 0 to 50000 | 176 (33.0%) | 20 (17.2%) | 66 (28.4%) | 90 (48.6%) |  |
| 50001 to 100000 | 70 (13.1%) | 14 (12.1%) | 32 (13.8%) | 24 (13.0%) |  |
| 100001 to 150000 | 32 (6.0%) | 7 (6.0%) | 14 (6.0%) | 11 (5.9%) |  |
| 150001 to 200000 | 45 (8.4%) | 13 (11.2%) | 22 (9.5%) | 10 (5.4%) |  |
| >200000 | 207 (38.8%) | 61 (52.6%) | 97 (41.8%) | 49 (26.5%) |  |
| **Household Income, missing: 3** |  |  |  |  | **<0.010 \*\*** |
| 0 to 50000 | 47 (8.8%) | 3 (2.6%) | 14 (6.0%) | 30 (16.2%) |  |
| 50001 to 100000 | 56 (10.5%) | 8 (6.9%) | 18 (7.8%) | 30 (16.2%) |  |
| 100001 to 150000 | 31 (5.8%) | 6 (5.2%) | 5 (2.2%) | 20 (10.8%) |  |
| 150001 to 200000 | 63 (11.8%) | 9 (7.8%) | 35 (15.1%) | 19 (10.3%) |  |
| >200000 | 333 (62.5%) | 89 (76.7%) | 159 (68.5%) | 85 (45.9%) |  |
| **Number of People in Household, *missing: 1*** | 3.62 (1.79) | 3.55 (1.55) | 3.57 (1.93) | 3.71 (1.75) | 0.655 |

**Caption:** Unadjusted associations by Pregnancy Status. Data are shown as <sup>1</sup>N (%) for categorical variables and mean (SD) for continuous variables. Continuous data were compared using Analysis of Variance (ANOVA) type 3 sum of squares. Categorical data were compared using Pearson's chi-square test. The symbol (\*\*\*) denotes that a P value is statistically significant at the $\alpha = 0.001$ level. The symbol (\*\*) denotes that a P value is statistically significant at the $\alpha = 0.01$ level. The symbol (\*) denotes that a P value is statistically significant at the $\alpha = 0.05$ level. For the variable *Tribe*, the sub-category of *Other African* primarily includes the following tribes: Iraq, Mmeru, Muha, among others not mentioned.

For ACDP, the majority (56.9%) of pregnant women reported complete abstinence, with the remainder drinking 1 to 2 times per week. Women not of childbearing age had higher rates of alcohol use, with 17.9% drinking 3 to 4 times per week. Of the pregnant women who drank, 32.8% consumed 1 to 2 standard drinks in a single sitting and 9.5% consumed 3 to 4 standard drinks in a sitting. Non-pregnant women were more likely to drink 3 to 4 standard drinks in a single sitting (16.8% in non-pregnant, young women and 13.5% in non-pregnant, older women). Following increased alcohol use, more older, non-pregnant women (55.1%) reported attempts to quit alcohol than childbearing-aged non-pregnant women (41.4%), and pregnant women (33.6%). Few childbearing-aged non-pregnant women (6.5%) and non-childbearing-aged non-pregnant women (5.4%) sought treatment for their alcohol use. Younger, non-pregnant women had the highest mean AUDIT score (2.94), compared to older non-pregnant women (2.51) and pregnant women (1.70) (Table 2).

**Table 2:** Alcohol Use Practices, Consequences and Depression Status by Pregnancy Status.

| *Alcohol Use Practices by Pregnancy Status* | *Overall*<br>*(n = 533)* | *Pregnant*<br>*(n = 116)* | *Not Pregnant,*<br>*Child-bearing*<br>*(n = 232)* | *Not Pregnant, Not*<br>*Child-bearing*<br>*(n = 185)* | *P Value* |
| --- | --- | --- | --- | --- | --- |
| **Drinking Frequency, (n,%), missing: 2** |  |  |  |  | **< 0.001 \*\*\*** |
| 0 times/week | 243 (45.6%) | 66 (56.9%) | 115 (49.6%) | 62 (33.5%) |  |
| 1 to 2 times/week | 227 (42.6%) | 45 (38.8%) | 93 (40.1%) | 89 (48.1%) |  |
| 3 to 4 times/week | 43 (8.1%) | 4 (3.4%) | 20 (8.6%) | 19 (10.3%) |  |
| 5 to 6 times/week | 4 (0.8%) | 0 (0%) | 2 (0.9%) | 2 (1.1%) |  |
| Everyday | 14 (2.6%) | 0 (0%) | 2 (0.9%) | 12 (6.5%) |  |
| **Drinking Quantity (# standard drinks per sitting), (n,%) missing: 1** |  |  |  |  | **< 0.001 \*\*\*** |
| 0 standard drinks | 244 (45.8%) | 66 (56.9%) | 115 (49.6%) | 63 (34.1%) |  |
| 1 to 2 standard drinks | 202 (37.9%) | 38 (32.8%) | 72 (31.0%) | 92 (49.7%) |  |
| 3 to 4 standard drinks | 75 (14.1%) | 11 (9.5%) | 39 (16.8%) | 25 (13.5%) |  |
| 5 to 6 standard drinks | 10 (1.9%) | 0 (0%) | 5 (2.2%) | 5 (2.7%) |  |
| >6 standard drinks | 1 (0.2%) | 0 (0%) | 1 (0.4%) | 0 (0%) |  |
| **Weekly Alcohol Expenses (TZS), (n,%), missing: 1** |  |  |  |  | **<0.01 \*\*** |
| 0 to 10000 | 460 (86.3%) | 98 (84.5%) | 190 (81.9%) | 172 (93.0%) |  |
| 10001 to 50000 | 69 (12.9%) | 17 (14.7%) | 39 (16.8%) | 13 (7.0%) |  |
| 50001 to 100000 | 3 (0.6%) | 0 (0%) | 3 (1.3%) | 0 (0%) |  |
| **Alcohol Preferences, (n,%), missing: 1** |  |  |  |  | **<0.001 \*\*\*** |
| Wine | 67 (12.6%) | 24 (20.7%) | 37 (15.9%) | 6 (3.2%) |  |
| Light Beer | 38 (7.1%) | 8 (6.9%) | 24 (10.3%) | 6 (3.2%) |  |
| Beer | 106 (19.9%) | 15 (12.9%) | 39 (16.8%) | 52 (28.1%) |  |
| Mbege (banana-based beer) | 67 (12.6%) | 2 (1.7%) | 13 (5.6%) | 52 (28.1%) |  |
| Other | 12 (2.4%) | 0 (0%) | 1 (0.4%) | 6 (3.2%) |  |
| None | 240 (45.0%) | 66 (56.9%) | 112 (48.3%) | 62 (33.5%) |  |
| **Attempted Quitting at any Time-Point, (n,%), missing: 4** |  |  |  |  | **< 0.01 \*\*** |
| No | 292 (54.8%) | 75 (64.7%) | 135 (58.2%) | 82 (44.3%) |  |
| Yes | 237 (44.5%) | 39 (33.6%) | 96 (41.4%) | 102 (55.1%) |  |
| **Sought Treatment for Alcohol Use, (n,%), missing: 4** |  |  |  |  | 0.063 |
| Yes | 25 (4.7%) | 0 (0%) | 15 (6.5%) | 10 (5.4%) |  |
| No | 504 (94.6%) | 116 (100%) | 215 (92.7%) | 173 (93.5%) |  |
| **AUDIT Score, (mean, SD), missing: 0** | 2.55 (4.39) | 1.70 (3.28) | 2.94 (4.79) | 2.51 (4.36) | **0.045 \*** |
| **HHD Status, (n,%), missing: 0** |  |  |  |  | 0.092 |
| Yes | 69 (12.9%) | 10 (8.6%) | 38 (16.4%) | 21 (11.4%) |  |
| No | 464 (87.1%) | 106 (91.4%) | 194 (83.6%) | 164 (88.6%) |  |
| **DrInC Score, (mean, SD), missing: 0** | 5.44 (12.0) | 3.15 (8.97) | 6.73 (12.6) | 5.25 (12.7) | **0.030 \*** |
| **PHQ9 Score, (mean, SD), missing: 0** | 6.10 (4.87) | 4.71 (3.12) | 5.85 (4.80) | 7.29 (5.55) | **<0.001 \*\*\*** |
| **Depression (PHQ-9 ≥9), (n,%), missing: 0** |  |  |  |  | **<0.001 \*\*\*** |
| Yes | 127 (23.8%) | 14 (12.1%) | 55 (23.7%) | 58 (31.4%) |  |
| No | 406 (76.2%) | 102 (87.9%) | 177 (76.3%) | 127 (68.6%) |  |
| **Sought Psychiatric Treatment, (n,%), missing: 3** |  |  |  |  | 0.094 |
| Yes | 51 (9.4%) | 11 (9.5%) | 29 (12.5%) | 11 (5.9%) |  |
| No | 487 (90.0%) | 105 (90.5%) | 200 (86.2%) | 174 (94.1%) |  |

**Caption:** Unadjusted associations with Alcohol Use Practices by Pregnancy Status. Data are shown as N (%) for categorical variables and mean (SD) for continuous variables. Continuous data were compared using Analysis of Variance (ANOVA) test with type 3 sum of squares. Categorical data were compared using Pearson's chi-square test. The symbol (\*\*\*) denotes that a P value is statistically significant at the $\alpha = 0.001$ level. The symbol (\*\*) denotes that a P value is statistically significant at the $\alpha = 0.01$ level. The symbol (\*) denotes that a P value is statistically significant at the $\alpha = 0.05$ level. For the variable *Alcohol Preferences*, the sub-category of *Other* primarily includes the following: Liquor/Spirits, Daddi, Ulanzi, Piwa, Gongo, Changaa, among others not mentioned.

Compared to pregnant women, younger non-pregnant women reported experiencing significantly more alcohol-related consequences, with an average DrInC score of 6.73, followed by 5.25 in older non-pregnant females, and 3.15 in pregnant females (Table 2). Depression severity was also highest amongst older non-pregnant women, with an average PHQ-9 score of 7.25, followed by 5.85 in younger non-pregnant women and 4.71 in pregnant women (Table 2). Older non-pregnant women (31.4%) were much more likely to be classified as depressed, defined as a PHQ-9 score greater than or equal to 9 points. Similar proportions of all female subgroups report seeking treatment for psychiatric disease, with rates ranging from 5.9% to 12.5% (Table 2).

Most (70.7%) women claimed that consuming 0 alcoholic drinks was the “safe amount” and reported ACDP is unacceptable throughout all stages of pregnancy. In contrast, over a quarter of older non-pregnant women (25.9%) believed that 1 to 2 standard drinks was a “safe amount” of alcohol while pregnant. (Table 3).

**Table 3.**
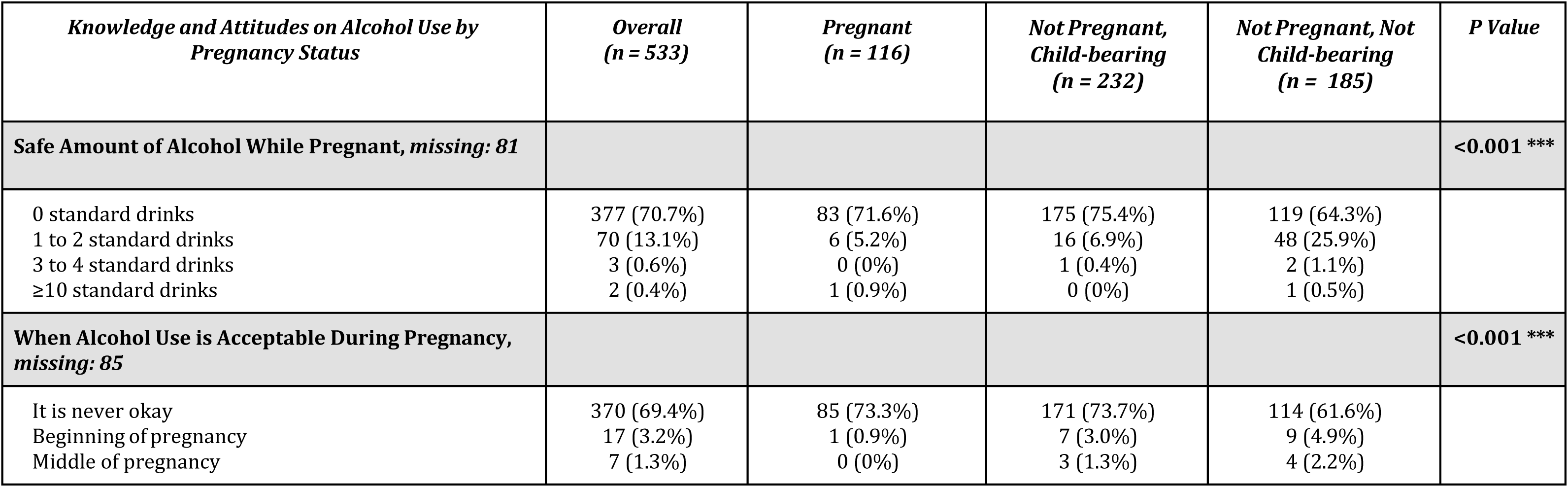
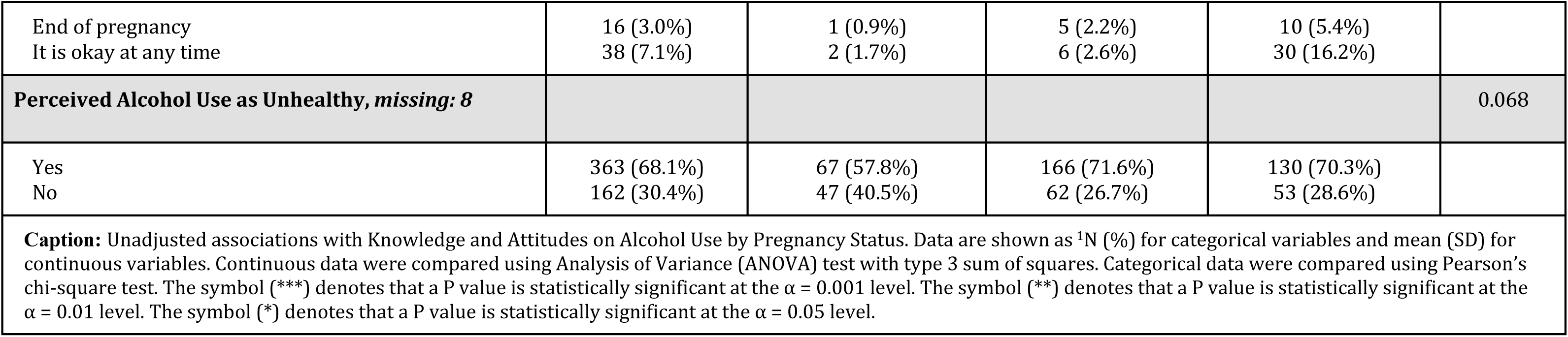
Survey-Reported Knowledge and Attitudes on Alcohol Use by Pregnancy Status.

| *Knowledge and Attitudes on Alcohol Use by Pregnancy Status* | *Overall*<br>*(n = 533)* | *Pregnant*<br>*(n = 116)* | *Not Pregnant,*<br>*Child-bearing*<br>*(n = 232)* | *Not Pregnant, Not*<br>*Child-bearing*<br>*(n = 185)* | *P Value* |
| --- | --- | --- | --- | --- | --- |
| **Safe Amount of Alcohol While Pregnant, missing: 81** |  |  |  |  | **<0.001 \*\*\*** |
| 0 standard drinks | 377 (70.7%) | 83 (71.6%) | 175 (75.4%) | 119 (64.3%) |  |
| 1 to 2 standard drinks | 70 (13.1%) | 6 (5.2%) | 16 (6.9%) | 48 (25.9%) |  |
| 3 to 4 standard drinks | 3 (0.6%) | 0 (0%) | 1 (0.4%) | 2 (1.1%) |  |
| ≥10 standard drinks | 2 (0.4%) | 1 (0.9%) | 0 (0%) | 1 (0.5%) |  |
| **When Alcohol Use is Acceptable During Pregnancy, missing: 85** |  |  |  |  | **<0.001 \*\*\*** |
| It is never okay | 370 (69.4%) | 85 (73.3%) | 171 (73.7%) | 114 (61.6%) |  |
| Beginning of pregnancy | 17 (3.2%) | 1 (0.9%) | 7 (3.0%) | 9 (4.9%) |  |
| Middle of pregnancy | 7 (1.3%) | 0 (0%) | 3 (1.3%) | 4 (2.2%) |  |
| End of pregnancy<br>It is okay at any time | 16 (3.0%)<br>38 (7.1%) | 1 (0.9%)<br>2 (1.7%) | 5 (2.2%)<br>6 (2.6%) | 10 (5.4%)<br>30 (16.2%) |  |
| **Perceived Alcohol Use as Unhealthy, *missing: 8*** |  |  |  |  | 0.068 |
| Yes<br>No | 363 (68.1%)<br>162 (30.4%) | 67 (57.8%)<br>47 (40.5%) | 166 (71.6%)<br>62 (26.7%) | 130 (70.3%)<br>53 (28.6%) |  |
| **Caption:** Unadjusted associations with Knowledge and Attitudes on Alcohol Use by Pregnancy Status. Data are shown as <sup>1</sup>N (%) for categorical variables and mean (SD) for continuous variables. Continuous data were compared using Analysis of Variance (ANOVA) test with type 3 sum of squares. Categorical data were compared using Pearson's chi-square test. The symbol (***) denotes that a P value is statistically significant at the $\alpha = 0.001$ level. The symbol (**) denotes that a P value is statistically significant at the $\alpha = 0.01$ level. The symbol (*) denotes that a P value is statistically significant at the $\alpha = 0.05$ level. | | | | | |

**Table 4:**
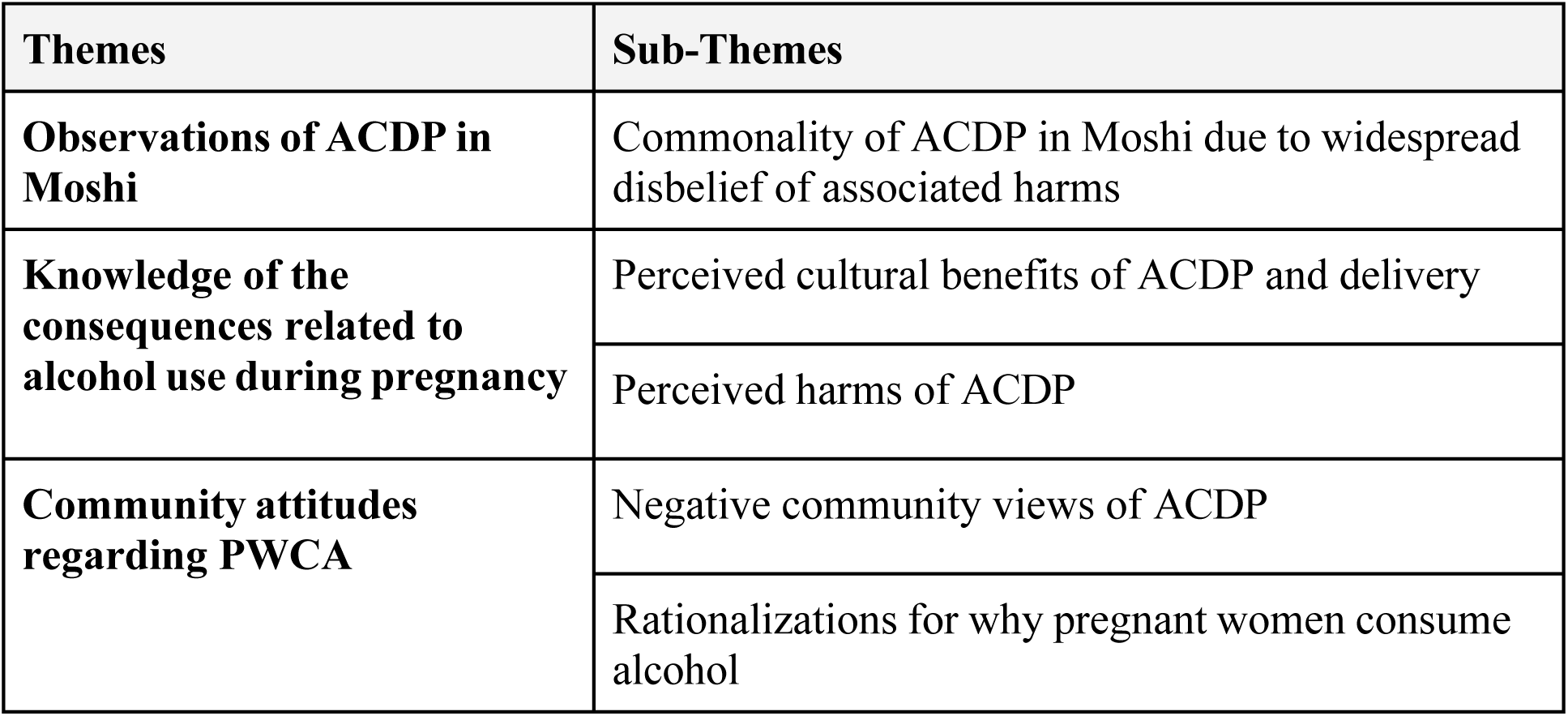
Qualitative Themes and Sub-Themes Regarding Alcohol Use and Pregnancy.

### Qualitative Results

Ten women and nine men completed an IDI. Out of the 10 women who participated in the IDIs, two had never been pregnant, two were pregnant at the time of the interview, and six had been pregnant in the past. Quotes drawn from these 10 women were labelled according to pregnancy status, as either ‘Current Pregnancy’, ‘Past Pregnancy’, or ‘Never Pregnant’. Of the 9 men who participated in IDIs, all had either witnessed PWCA in their local communities or had cohabited with their pregnant female partner in a registered or unregistered marriage.

IDI participants ranged from 20 to 70 years old, with marital statuses including single or never married, married, divorced, and widowed and education statuses from primary to college level. Alcohol consumption patterns ranged from complete abstinence to frequent consumption, such as 3 to 4 bottles per sitting nearly 5 to 6 times per week. Three themes were inductively and deductively identified from the 19 interviews: (1) Observation of ACDP in Moshi, including its commonality, (2) Knowledge of the consequences related to alcohol use during pregnancy, such as the influence of cultural beliefs and perceived harms, and (3) Community attitudes regarding PWCA.

### Observations of ACDP in Moshi

#### Commonality of ACDP in Moshi due to widespread disbelief of associated harms

Over half of the IDI participants noted that it was “*quite common to see a pregnant woman drinking alcohol”* (IDI #6, Female, Past Pregnancy) in Moshi, with almost half of female interviewees reporting alcohol consumption themselves during a current, or past pregnancy. One participant recalled:

> *“When I was pregnant, I used to go to the clinic and being told not to drink alcohol but when I returned home, I continued to drink alcohol as usual” (IDI #6, Female, Past Pregnancy)*

Another female respondent shared her observations of PWCA, which shaped her own alcohol use.

> *“I have seen many [pregnant women] who use alcohol, some of them even drink a lot of beer bottles but I have never witnessed any of them getting any bad effect even after delivery … Drinking wine helps me with my health, such as in food digestion process. That’s why I decided to continue drinking even when pregnant … I will drink just 2 glasses of wine per week. It’s such a little amount that am sure cannot bring harm to my baby.” (IDI #19, Female, Current Pregnancy)*

Several other participants witnessed a lack of observed adverse consequences when consuming alcohol, which appeared to contribute to the commonality of this practice in Moshi. This sentiment is accurately summarized by one participant who highlights that a mother gave birth to a healthy baby after consuming alcohol throughout her pregnancy.

> *“There was a woman; when she was pregnant, she drank so much alcohol. One day, the baby turned up, we brought her here to KCMC hospital. She was drunk and she was told it was still not the time to give birth. She returned home and continued drinking alcohol until her delivery, yet she still gave birth to a healthy baby.” (IDI #6, Female, Past Pregnancy)*

Another participant had a similar sentiment, which describes how a mother gave birth to an unhealthy baby while not drinking during pregnancy and a healthy baby while drinking during pregnancy.

> *“During the first pregnancy, she did everything … she had to do, she even stopped drinking alcohol but the baby was still born with problems. After giving birth to the first child, she started to drink alcohol, even when she was pregnant with the second child… She refused to stop drinking alcohol until she gave birth…and she had a normal child with no problems” (IDI #9, Female, Past Pregnancy)*

These results highlight the commonality of alcohol use during pregnancy in Moshi Tanzania due to unobserved harms with the practice. In contrast to these excerpts, the remaining IDI participants stated that ACDP was “*not a common practice”* (IDI #11, Male) in Moshi due to community sensitization to alcohol-related harms on the mother and fetus, whereby “*people are aware of the effects of alcohol during pregnancy”* (IDI #11, Male) and “*they understand the effects of alcohol*” (IDI #5, Male)

### Knowledge of consequences related to alcohol use during pregnancy

#### Perceived cultural benefits of ACDP and delivery

Cultural beliefs surrounding ACDP had both tribal and cultural origins. For example, nearly a quarter of participants highlighted that members of the Chagga tribe encouraged moderate alcohol consumption before, during, and after delivery. These norms encouraged pregnant women to “*sip a small amount of alcohol,*” (IDI #8, Male), as this was thought to provide benefit to the pregnant female and unborn child. Specifically, it was thought that alcohol would “*help the baby [to] grow big and become active*,” (IDI #11, Male) with participants encouraging alcohol use “*at the middle of pregnancy*” because “*at that stage, the baby should grow and be strong before delivery*” (IDI #8, Male)

Participants likewise highlighted numerous perceived benefits of alcohol use during labor, including providing the mother with “*increased power*” (IDI #7, Female, Past Pregnancy) to push the baby through the birth canal, alleviating stress, “*[reducing] labor pain”* (IDI #14, Male) and “*abdominal cramps*” (IDI #12, Male), and “*increase milk production for my baby to breastfeed*” (IDI #7, Female, Past Pregnancy). One female interviewee elaborated on milk production, recounting that:

> *“There is a notion where I live that taking alcohol during and after delivery increases milk production and eventually the baby gets enough milk to breastfeed…after my third delivery where my grandmother insisted for me to take it in order to increase milk production for my baby to breastfeed.” (IDI #7, Female, Past Pregnancy)*

These beliefs, however, were not characteristic of all parts of Moshi. One participant highlights a distinct discrepancy between village views and town views concerning PWCA, explaining that ACDP is prevalent and accepted only in certain cultural contexts:

> *“The community views [PWCA] differently. If you ask people from the village, they would say its right, but in town, because people are aware of the effect of alcohol, you may find few of them who drink …” (IDI #14, Male)*

These results highlight the underlying cultural beliefs on the benefits of consuming alcohol during pregnancy that may be influencing consumption patterns.

#### Perceived harms of ACDP

While cultural influence may support the use of alcohol during pregnancy, many participants noted substantial harms of ACDP on the developing fetus, including premature birth, fetal anomalies, and nutritional deficiencies. Additionally, respondents also expressed these harms extend to the mother, including a detrimental impact on mental health and motor functioning. One respondent notes:

> *“The baby may be born with some abnormalities such as blindness, mental problems or born prematurely. I have seen one female who was drinking during pregnancy, and she end up delivering a premature baby.” (IDI #11, Male)*

Several participants also reported that ACDP contributed to the development of blindness, neurological, cognitive, and motor disabilities. Of equal concern, a handful of participants suggested that alcohol-induced appetite loss in expectant mothers could lead to nutritional deficiencies hindering proper fetal development.

> *“Alcohol sometimes causes someone to lose appetite, it depends on the type of alcohol you drink. You may drink alcohol and fail to eat properly, and harm the baby or the mother can get anemia.” (IDI #9, Female, Past Pregnancy)*

The impact of alcohol use was seen to extend into the postnatal period. About a quarter of interviewees cautioned against alcohol use during breastfeeding, noting that “*it will have impact on child’s mental and motor development and eventually restricts normal and proper growth of that child*” (IDI #7, Female, Past Pregnancy).

These risks extended to the mother as well, with a fifth of interviewees asserting that ACDP can have a detrimental impact on female health, including engaging in unsafe sexual behaviors or disrupting motor function, where an intoxicated pregnant women may “*fall down and injure the stomach…[with] a possibility of terminating pregnancy*” (IDI #14, Male). As one interviewee explained:

> *“In general, alcohol consumption during pregnancy is not good for a pregnant woman to drink too much alcohol. When got drunkard she can do unprotected sex with someone she does not know and put herself at risk of sexual transmitted infections or AIDS during pregnancy.” (IDI #9, Female, Past Pregnancy)*

Finally, a few interviewees emphasized the importance of adequate education on the detrimental effects of ACDP, noting that “*[Women] are always ready to try their best when they are given right knowledge and education about the consequences of alcohol to themselves until delivery time and to their unborn babies*” (IDI #16, Female, Never Pregnant).

### Community attitudes regarding PWCA

#### Negative community views of ACDP

Generally, community perspectives cast PWCA in a negative light. Most IDI participants claimed that ACDP was prohibited and engaging in such behaviors would quickly attract social disapproval. To avoid censure, one respondents specifically mentioned that PWCA tended to *“drink privately”* (IDI #8, Male). Interviewees described that PWCA are viewed as incapable of suppressing alcoholic urges and of making sacrifices for the sake of her child. This shared community attitude was epitomized by the language used by respondents to describe PWCA who “*does not wish good for her baby”* (IDI #3, Female, Never Pregnant). Participants claimed that the community views PWCA as “*foolish*” (IDI #10, Female, Never Pregnant) and “*taboo,”* (IDI #9, Female, Past Pregnancy) whereby alcohol consumption while pregnant is “*careless*” (IDI #3, Female, Never Pregnant). Additionally, one participant stated, “*The community perceive them as ‘risk taker’,’don’t care’ they can even be isolated in the community,*” (IDI #4, Male) thereby demonstrating the profound influence of these adverse community perceptions on PWCA.

Several participants compared PWCA to non-pregnant women who consume alcohol, implying that PWCA are viewed particularly poorly. Specifically, participants posited that since pregnant women actively “*agreed to conceive,*” (IDI #9, Female, Past Pregnancy) they and their partner bore the responsibility of ensuring the well-being and health of themselves, and of the unborn child. For example, participants remarked that “*whether she is educated or not education, a pregnant woman drinking alcohol will seem it is it so wrong*” (IDI #9, Female, Past Pregnancy) and when a woman drinks while pregnant, “*[the community] normally does some traditional ritual to her*” (IDI #11, Male).

#### Rationalizations for why pregnant women consume alcohol

Several factors, collectively, were rationalized by participants to cause a pregnant woman to either increase, decrease, or altogether abstain from alcohol consumption during pregnancy. First, nearly half of the interviewees associated ACDP with an inability to stop consumption due to pre-existing alcohol dependency, with stress that occurred before a woman was pregnant was hypothesized to underlie pre–pregnancy alcohol use.

> *“Some of them were good drinkers even before pregnancy therefore its not easy for them to stop abruptly during pregnancy… I think it’s just because she was already used to taking alcohol regularly when she wasn’t pregnant and she failed to stop using.” (IDI #3, Female, Never Pregnant)*

> *“For sure I have no idea of why she takes alcohol at that [perinatal] period, but I know she has been using all that since long time ago, at some point she had family conflicts with her husband which may be one of the reasons” (IDI #7, Female, Past Pregnancy)*

Moreover, participants held the belief that pregnant women are likely to increase alcohol consumption if they experienced an unexpected pregnancy while unmarried. Such situations were thought to induce stress and societal pressures for these expectant mothers, as elucidated by one participant’s remark:

> *“We have one problem in our community, most men run away once they have [kids]. So due to stress, this women start or increase the amount of drinking alcohol” (IDI #18, Male)*

Second, over a quarter of participants believed that pregnancy hormones may cause a *“strong desire and cravings”* (IDI #3, Female, Never Pregnant) for certain foods and drinks, including alcohol. For example, one participant explained how pregnancy cravings may influence alcohol use:

> *“There are quite a number of pregnant women who take alcohol. Depending on their desires and pregnancy cravings, some drink a lot, others due to pregnancy don’t even want to smell [alcohol], others remain using as they were before” (IDI #7, Female, Past Pregnancy)*

While generally condoned, continued reasons for ACDP remain complex and multi-faceted.

## 4 DISCUSSION

This study aimed to explore perceptions and assess current rates of alcohol use among pregnant women in Moshi. To the best of our knowledge, this is one of the first studies using a mixed-methods approach to closely examine community attitudes around ACDP in Moshi, Tanzania. Our analysis found that rates of alcohol use among pregnant women in Moshi remain high, with the biggest predictor of ACDP being pre-pregnancy alcohol use. While overall, the community held negative views around cultural beliefs and believed women should not drink at any point in their pregnancy, some sociocultural beliefs encouraged ACDP. The high rates of alcohol use and complex factors influencing use point to the need for robust community-wide health messaging coupled with patient-level, pre-pregnancy education and holistic support in order to reduce this generational harm.

ACDP in Moshi remains notably elevated. We found that 42% of pregnant women attending KCMC’s ED and RHC consume alcohol weekly, drinking 1 to 2 standard drinks of alcohol per sitting, with previous alcohol use being most associated with use during pregnancy. Our qualitative results supported the commonality of ACDP, with this behavior noted to be frequently observed in local communities, spurred by a lack of observed consequences. These rates are significantly higher than previous, nearby studies that found ACDP to be 20% in Northern Tanzania and 15% in neighboring Dodoma ^14,16^. These findings, particularly the ubiquitousness of ACDP, are cause for concern given existing recommendations that pregnant women should abstain from alcohol entirely ^13^. Populations exhibiting high rates of alcohol consumption are typically associated with a high prevalence of FASD ^9–12^; however, within Tanzania, like most of SSA, the true prevalence of FASD is not well understood and corresponding literature is rare^42^. FASD and other alcohol-related congenital anomalies typically develop early in pregnancy, often before women realize they are pregnant. Importantly, alcohol use behaviors before pregnancy were identified as predicting ACDP as revealed in IDIs, with several participants noting pre-pregnancy alcohol habits to be the primary reason for continued alcohol use. This is especially important to consider as young women who are currently not, but may become pregnant, exhibited higher levels of alcohol use (AUDIT = 2.94) and alcohol-related consequences (DrInC = 6.73), than other female subgroups. Previous studies have identified pre-pregnancy alcohol use among young women to be a significant predictor of alcohol use during pregnancy ^14,43^, with literature noting that the minimization of these habits are key in reducing ACDP ^14^.

In both our quantitative and qualitative analysis, ACDP was generally censured, however, sociocultural and community observations and beliefs underlied continued alcohol use during pregnancy. Survey data revealed that 70% of participants found any amount of alcohol to be unsafe at any point during pregnancy, a finding qualitatively supported by the general negative community views of PWCA and notable mentioned harms of this behavior. The observed societal stigma towards women who drink, regardless of pregnancy, warrants further investigation to determine if this stigma is more profound in pregnancy, or applies generally to all women. Aside from previous alcohol use, IDI responses provided a more nuanced understanding of why ACDP continued within Moshi, mentioning stress, addiction, and observations of other women who drank and gave birth to healthy children. It is imperative to note that accessing mental health support is challenging in Moshi, Tanzania, particularly for women; only a small percentage of women (9.4%) sought psychiatric treatment programs, despite a high prevalence of depression (23.8%). Marlow et al. similarly found cultural beliefs played a role in alcohol use among pregnant women in Lesotho and echoed the sentiment of alcohol as protective for the unborn baby^44^. Previous literature has also found stress and depression to be highly prevalent in women from SSA both during and after pregnancy ^45–47^, with stress also linked to increased alcohol consumption both in this region and worldwide ^48–51^. The influence of stress and addiction underlying ACDP points to a need for future programming to incorporate components of mental healthcare, including improved availability and access to mental healthcare, and the minimization of alcohol use among both pregnant and non-pregnant women of childbearing age ^49,52–55^.

These factors illustrate the necessity of increased community-wide health messaging centered around the dangers of ACDP, coupled with patient-level, pre-pregnancy education and holistic support for women of childbearing age. For example, our analysis found that persistent beliefs, painting ACDP as beneficial exist, illustrating that a gap remains in community education and practice. This call to action joins other literature, which similarly has identified high rates of alcohol use during pregnancy in SSA, to put greater focus on developing and implementing behavioral interventions ^56^. Interventions to reduce ACDP have been tried and tested worldwide, with formats being primarily medical-, psychosocial-, and education-centered ^57^. The most effective interventions for pregnant women were psychosocial-based, specifically brief interventions (BIs), which is a focused, motivational interview delivered directly to the patient and can be readily incorporated into prenatal care visits ^58^. This intervention type may be especially useful in Moshi, as BIs have been shown to be particularly effective in resource-constrained settings, requiring little cost to operate and can be performed by lay healthcare workers ^59^. Future programming in this region should incorporate both increased community messaging while also delivering effective education and support directly to expectant mothers.

Local policy efforts should be aware of the high rates of alcohol use still present in Moshi and re-focus health priorities on mitigating this preventable harm. Additionally, to better inform the creation of appropriate interventions and programming, future studies should investigate the extent of current alcohol-related fetal harm in Tanzania to ensure the most impactful prevention strategies are utilized. Through these strategies, alcohol-related harm can be mitigated, with improved health outcomes for present and future generations.

### Strengths and Limitations

The primary limitations of this study include selection bias, as the population was recruited solely from one health facility in Moshi, which may not adequately represent the broader Moshi community, and recall bias, as alcohol use behaviors were self-reported through surveys. That being said, this one hospital evaluation will serve as a contextual framework for an implementation study and further literature about barriers to alcohol harm reduction in low resource settings. Additionally, it is important to acknowledge the uncertainty surrounding the observed prevalence of pregnant women who consume alcohol (42.5%) given the limited size of our sample of pregnant women (n = 116).

## 5 CONCLUSION

Our study highlights both current high rates of alcohol consumption among pregnant women in Moshi, a behavior shaped in part by cultural beliefs and practices, alongside negative community attitudes towards these individuals. Stress coping, existing alcohol dependency, and pregnancy-related cravings constitute additional motivations for alcohol consumption among pregnant women. These findings stress the need for future supportive, education-based programming for young women of childbearing age. More holistic pregnancy-related healthcare services and infrastructure in this region may be the solution to curbing alcohol use and improving health outcomes within the Moshi community.

## Supporting information

S1_Checklist

S2_Checklist

S1_Questionnaire

## Data Availability

Data are only available upon reasonable request, as participants did not consent to public data transfer and requires a written agreement approved by Kilimanjaro Christian Medical Centre Ethics Committee and the National Institute for Medical Research (Tanzania). Data inquiries can be sent to Gwamaka W. Nelson at.

## Author Contributions

Conceptualization: AP, KW, CAS, SR

Methodology: AP, KW, SR, CAS

Formal analysis and investigation: KW, CAS, BTM, BM, SR, FS

Data Collection: MM, YS, JK, AP

Writing - original draft preparation: KW, AP, MB, SR

Writing - review and edition: KW, AP, FS, SR, BM, BTM, CAS

Funding acquisition: AP, CAS

Supervision: CAS, SR, BTM

## Abbreviations

PWCA: Pregnant women who consume alcohol
ACDP: Alcohol consumption during pregnancy
SSA: Sub-Saharan Africa

## Supporting Information

1. **S1 File**
2. **S2 File**
3. **S3 File**
4. **S4 File**
5. **S1 Checklist**
6. **S2 Checklist**
7. **S1 Questionnaire**

