## Supplementary material for "The Burden of Generational Harm due to Alcohol use in Tanzania: a mixed method study of pregnant women": S1_file

DukeHealth
Institutional Review Board

### DUHS INSTITUTIONAL REVIEW BOARD NOTIFICATION OF STUDY APPROVAL

**Protocol ID:** Pro00107861
**Reference ID:** Pro00107861-INIT-1.0
**Principal Investigator:** Catherine Staton
**Protocol Title:** Differences in Alcohol Use Behaviors between Men and Women in Moshi, Tanzania: A Mixed Methods Study

**Sponsor/Funding Source(s):** Duke Global Health Institute(1859)
**Federal Funding Agency ID:**

The Duke University Health System Institutional Review Board for Clinical Investigations has conducted the following activity on the study cited above:
**Activity:** New Study

**Review Type:** Expedited

**Review Date:** 06/03/2021

**Issue Date:** 06/04/2021

**Expiration Date:** 06/03/2023

DUHS IRB approval encompasses the following specific components of the study:
**Protocol version/date:**
**DUHS IRB Application version:** v1.1
**Consent form reference date:** 06/04/2021 - Consent Form (Swahili)
**Investigator Brochure, version/date:**
**Pediatric Risk Category:**
**Other:**
- research_ethics_certification_tanzania_team
- certificate of translation
- consent_form_english
- Interview Questions (English and Swahili)
- AUDIT_questionnaire

The DUHS IRB has determined the specific components above to be in compliance with all applicable Health Insurance Portability and Accountability Act ("HIPAA") regulations.

This study qualifies for transition to the revised Common Rule effective January 21, 2019 and the elimination of annual review by the IRB. Please note that this study will only require completion of an abbreviated report on the study's progress over the last 2 years beginning on the 2-year required check-in date cited above.

You must complete a 2-year check-in form in iRIS, or close the study before the 2-year required check-in date. The 2-year check-in submissions must be received by the DUHS IRB office 60 to 45 days prior to the 2-year required check-in date above.

No change to the protocol, consent form or other approved document may be implemented without first obtaining IRB approval for the change. Any proposed change must be submitted as an amendment. Safety reports must continue to be submitted according to IRB policy

The Duke University Health System Institutional Review Board for Clinical Investigations (DUHS IRB), is duly constituted, fulfilling all requirements for diversity, and has written procedures for initial and continuing review of human research protocols. The DUHS IRB complies with all U.S. regulatory requirements related to the protection of human research participants. Specifically, the DUHS IRB complies with 45CFR46, 21CFR50, 21CFR56, 21CFR312, 21CFR812, and 45CFR164.508-514. In addition, the DUHS IRB complies with the Guidelines of the International Conference on Harmonization to the extent required by the U. S. Food and Drug Administration.

DUHS Institutional Review Board
2424 Erwin Rd | Suite 405 | Durham, NC | 919.668.5111
Federalwide Assurance No: FWA 00009025
