## Supplementary material for "The Burden of Generational Harm due to Alcohol use in Tanzania: a mixed method study of pregnant women": S2_file

CRERC 07

KILIMANJARO CHRISTIAN MEDICAL UNIVERSITY COLLEGE

*(A Constituent College of Tumaini University Makumira)*

**P. O. Box 2240, MOSHI, Tanzania.**

**RESEARCH ETHICAL CLEARANCE CERTIFICATE**

**No. 2515**

**Research Proposal No.** 1297

**Study Title:** Differences in alcohol use behaviours between men and women in Moshi: A mixed method study

**Study Area** : KCMC referral hospital

**PIs Name** : Blandina Mmbaga

**Coinvestigators:** Catherine Staton, Joao Ricardo Nickenig Vissoci, Alena Pauley, Ashley Phillips

**Institution (s) :** Kilimanjaro Christian Medical University College

**The Proposal was approved by CRERC on :** 16th August, 2021

**Duration of Study :** One year

**From :** 16th August, 2021 to 15th August, 2022

**PROF. MRAMBA NYINDO**
**Chair – CRERC**

**PROF. EPHATA KAAYA**
**PROVOST - KCMU College**
