## Supplementary material for "The Burden of Generational Harm due to Alcohol use in Tanzania: a mixed method study of pregnant women": S3_file

### THE UNITED REPUBLIC OF TANZANIA

National Institute for Medical Research
3 Barack Obama Drive
P.O. Box 9653
11101 Dar es Salaam


Permanent Secretary (Health)
Ministry of Health, Community
Development, Gender, Elderly & Children
Government City Mtumba, Health Road
P.O. Box 743
40478 Dodoma

NIMR/HQ/R.8a/Vol. IX/3734

29th July 2021

Alena Pauley
Duke Global Health Institute
310 Trent Drive Rm 137, Durham NC, 27710
C/o Prof. Blandina Mmbaga
Kilimanjaro Christian Medical Centre
P O Box 2410
Moshi

**RE: ETHICAL CLEARANCE CERTIFICATE FOR CONDUCTING MEDICAL RESEARCH IN TANZANIA**

This is to certify that the research entitled: **Differences in alcohol use behaviors between men and women in Moshi, Tanzania: A mixed methods study (Pauley A. et al.)** whose local investigator is Prof. Blandina Mmbaga of Kilimanjaro Christian Medical Centre, has been granted ethical clearance to be conducted in Tanzania.

The Principal Investigator of the study must ensure that the following conditions are fulfilled:

1. Progress report is submitted to the Ministry of Health, Community Development, Gender, Elderly & Children and the National Institute for Medical Research, Regional and District Medical Officers after every six months.
2. Permission to publish the results is obtained from National Institute for Medical Research.
3. Copies of final publications are made available to the Ministry of Health, Community Development, Gender, Elderly & Children and the National Institute for Medical Research.
4. Any researcher, who contravenes or fails to comply with these conditions, shall be guilty of an offence and shall be liable on conviction to a fine as per NIMR Act No. 23 of 1979, PART III Section 10(2).
5. Sites: Kilimanjaro Christian Medical Centre.

Approval is valid for one year: 29th July 2021 to 28th July 2022.

**Name: Prof. Yunus Daud Mgaya**

**Signature**
**CHAIR PERSON**
**MEDICAL RESEARCH**
**COORDINATING COMMITTEE**

**Name: Dr. Aifello Wedson Sichalwe**

**Signature**
**CHIEF MEDICAL OFFICER**
**MINISTRY OF HEALTH, COMMUNITY DEVELOPMENT, GENDER, ELDERLY & CHILDREN**

CC: Director, Health Services-TAMISEMI, Dodoma.
RMO of Kilimanjaro region.
DMO/DED of Moshi district.
